# Gestational Age as a Predictor of Oxygen Needs and ABG Patterns in Preterm Neonates with RDS: Evidence from a Resource-Limited NICU in Pakistan

**DOI:** 10.1101/2025.10.04.25337307

**Authors:** Shoaib Mehmood, Ruhamah Yousaf

## Abstract

**Background:** Respiratory Distress Syndrome (RDS) remains a leading cause of morbidity and mortality among premature infants. Their immature lungs lack surfactant, making breathing difficult. These infants need close monitoring with arterial blood gas (ABG) checks to guide oxygen therapy. While gestational age is known to influence disease severity, locally relevant data on oxygen requirements and arterial blood gas (ABG) changes across gestational age groups remain scarce in Pakistan.

**Objective:** To evaluate gestational age–related variations in oxygen demand, ABG parameters, and respiratory support among preterm neonates with RDS, with the goal of identifying physiologic and management differences that may guide tailored interventions in low-resource NICU settings.

**Methodology:** This four-month cross-sectional study was conducted in the NICU of the University of Lahore Teaching Hospital, Pakistan. Sixty-five preterm neonates (28–36 weeks) with RDS were enrolled through consecutive sampling. Data on gestational age, oxygen requirement (FiO_2_), mode of respiratory support, and ABG parameters were collected before and after stabilization on respiratory support. Data were analysed in SPSS 27 using ANOVA, Kruskal–Wallis, Chi-square, paired t-tests, and Spearman’s correlation.

**Results:** Lower gestational age was linked to higher oxygen needs (FiO_2_ 83% at 28–30 weeks vs. 55% at 34–36 weeks, p < 0.001) and more invasive ventilation (60% vs. 10%, p = 0.011). ABG parameters correlated significantly with gestational age: earlier gestations had lower pH, higher PaCO_2_, and lower PaO_2_ and SaO_2_ (all p < 0.05), while HCO_3_^−^ showed no correlation (p = 0.316). Despite stabilization, SaO_2_ values remained below the target 90–94% range in all groups.

**Conclusion:** Gestational age is a key predictor of oxygen needs, ABG derangements, and intensity of respiratory support in preterm neonates with RDS. This first locally relevant evidence from Pakistan underscores the need for gestation-specific oxygen protocols to improve care in resource-limited NICUs and similar low-resource settings worldwide.

**Key Messages:** *What is already known on this topic:* Respiratory distress syndrome is a leading cause of illness and death in preterm babies. Oxygen therapy and arterial blood gas monitoring are central to care, but both under- and over-treatment can cause harm. Gestational age strongly influences oxygen needs and outcomes, yet data from local NICUs remain limited.

*What this study adds:* This study shows that babies born at lower gestational ages need higher FiO_2_ and more invasive support. Blood gas values such as pH, PaO_2_, PaCO_2_, and SaO_2_ vary significantly with gestational age, while HCO_3_^−^ does not. It also provides locally relevant evidence from Pakistan to guide neonatal care.

*How this study might affect research, practice or policy:* These findings highlight the need for gestation-specific oxygen therapy and monitoring strategies in NICUs. They support the development of standardized oxygen protocols in resource-limited settings. The results may also inform future research to improve outcomes and reduce preventable complications in preterm infants.

## INTRODUCTION

Respiratory distress syndrome (RDS) is a major cause of morbidity and mortality among preterm neonates. It results from pulmonary immaturity and insufficient surfactant, leading to alveolar collapse, impaired oxygenation, and respiratory failure. Infants born before 36 weeks are at greatest risk, although antenatal corticosteroids, surfactant therapy, and modern respiratory support have improved survival.(1, 2)

The pathophysiology of RDS involves alveolar collapse due to surfactant deficiency, which increases the work of breathing and reduces oxygen delivery to tissues. Many affected infants require supplemental oxygen, continuous positive airway pressure (CPAP), or invasive ventilation in severe cases. However, oxygen therapy carries risks. Hypoxia can result in organ dysfunction, while hyperoxia contributes to complications such as bronchopulmonary dysplasia (BPD), retinopathy of prematurity (ROP), and impaired neurodevelopment. Therefore, careful titration of FiO_2_ with close arterial blood gas (ABG) monitoring is essential to balance safety and efficacy.(3)

Gestational age strongly influences the severity of RDS and the level of support required. Extremely preterm infants often require higher FiO_2_, prolonged respiratory support, and demonstrate more deranged ABG values compared with more mature preterm.(4) Non-invasive strategies, particularly CPAP and high-flow nasal cannula (HFNC), are now widely preferred to reduce ventilator-induced lung injury.(5) In addition, less invasive surfactant administration techniques, such as thin-catheter delivery, have shown promise in reducing the need for intubation and improving outcomes.(6) In resource-limited settings where surfactant is scarce, CPAP has proven to be an effective first-line therapy and improves survival when initiated early.(7)

Management guidelines now recommend targeting oxygen saturation ranges between 90–95% to avoid both hypoxia and hyperoxia. This requires continuous FiO_2_ adjustment and ABG evaluation.(8) Evidence shows that very preterm infants (<28 weeks) not only require higher oxygen concentrations initially but also take longer to stabilize compared with later gestations.(9) ABG parameters, including pH, PaCO_2_, PaO_2_, HCO_3_^−^, and SaO_2_, remain critical indicators of respiratory and metabolic status, guiding therapy and predicting outcomes.(10)

Globally, preterm birth affects nearly 15 million infants each year, with a large proportion at risk of RDS. The burden is higher in low-resource countries where advanced interventions remain limited.(11) Despite improvements, oxygen-related complications continue to be a major concern.(12) Antenatal corticosteroids remain a cornerstone for promoting lung maturation and reducing disease severity.(13)

In Pakistan and other low- and middle-income countries, standardized oxygen protocols and advanced monitoring are not always available. There is limited local data on how oxygen requirements, ABG parameters, and respiratory support vary by gestational age in neonates with RDS. Addressing this evidence gap is essential to optimize oxygen therapy and reduce preventable complications.

Therefore, this study was conducted to compare oxygen requirements, ABG parameters, and respiratory support across gestational age groups in preterm neonates with RDS. By providing locally relevant data, it aims to support individualized, evidence-based neonatal care and improve outcomes in resource-constrained NICUs.

## METHODOLOGY

### Study Design and Setting

This hospital-based, observational analytical study with a comparative cross-sectional design was conducted in the Neonatal Intensive Care Unit (NICU) of University of Lahore Teaching Hospital, a tertiary care centre for preterm neonates with respiratory distress syndrome (RDS).

### Participants

Preterm neonates (<37 weeks gestation) diagnosed with RDS, admitted within 24 hours of birth, and requiring oxygen therapy with available arterial blood gas (ABG) analysis were included. Neonates with major congenital anomalies, sepsis, or birth asphyxia were excluded.

### Sample Size and Sampling

The sample size was calculated using a correlation-based formula:(14)

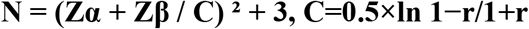

where *Z*_*α*_ = 1.96(α = 0.05), *Z*_*β*_ = 0.84(power = 80%), and r = 0.35 (expected correlation). Based on this, 62 neonates were required; 65 were enrolled using consecutive sampling to account for potential exclusions.(14, 15)

### Data Collection

Demographic details, gestational age, oxygen delivery device, FiO_2_, and ABG parameters (pH, PaO_2_, PaCO_2_, HCO_3_^−^, SaO_2_) were extracted from medical records using a structured questionnaire (attached). ABGs were collected just before starting respiratory support and after stabilization. Gestational age was the independent variable; FiO_2_, oxygen delivery device, and ABG parameters were dependent variables.

### Statistical Analysis

Continuous variables were summarized as mean ± SD and categorical variables as frequencies (%). Comparisons across gestational age groups used ANOVA or Kruskal-Wallis tests. Paired t-tests compared pre- and post-support values. Correlations between gestational age and oxygen/ABG parameters were assessed using Pearson or Spearman correlation. A p-value <0.05 was considered statistically significant.

### Ethical Approval and Informed Consent

This study was approved by the Departmental Review Board of The University of Lahore, Pakistan (Approval Letter No. Centre/Admin FAHS/195/25). The study was performed in accordance with the ethical standards of the institutional research committee and the principles outlined in the *Declaration of Helsinki*. Informed consent for participation was obtained verbally from the parents or legal guardians of all preterm neonates after explaining the purpose and procedures of the study. Verbal consent was chosen because most participants were approached during emergency NICU admissions, when obtaining written consent was not feasible. The verbal consent process was reviewed and approved by the Institutional Review Board.

### Patient and Public Involvement

Patients and/or the public were not involved in the design, conduct, reporting, or dissemination plans of this research.

## RESULTS

A total of 65 preterm neonates with respiratory distress syndrome (RDS) were included in the study. The distribution of participants by gestational age, sex, and baseline characteristics is summarized in Table 1. The majority of neonates (38.5%) were born at 31–33 weeks of gestation, followed by 28–30 weeks (30.8%) and 34–36 weeks (30.8%). Males constituted 56.9% of the study population, while 43.1% were females. Most neonates were of low birth weight (60.0%), and 30.8% were very low birth weight. Caesarean section was the predominant mode of delivery (73.8%), and 63.1% of neonates were inborn. Antenatal corticosteroids were administered in 78.5% of cases, and 63.1% received surfactant therapy. The 5-minute Apgar score was ≥7 in 60.0% of neonates, indicating satisfactory initial adaptation (Table 1).

**Table 1.**
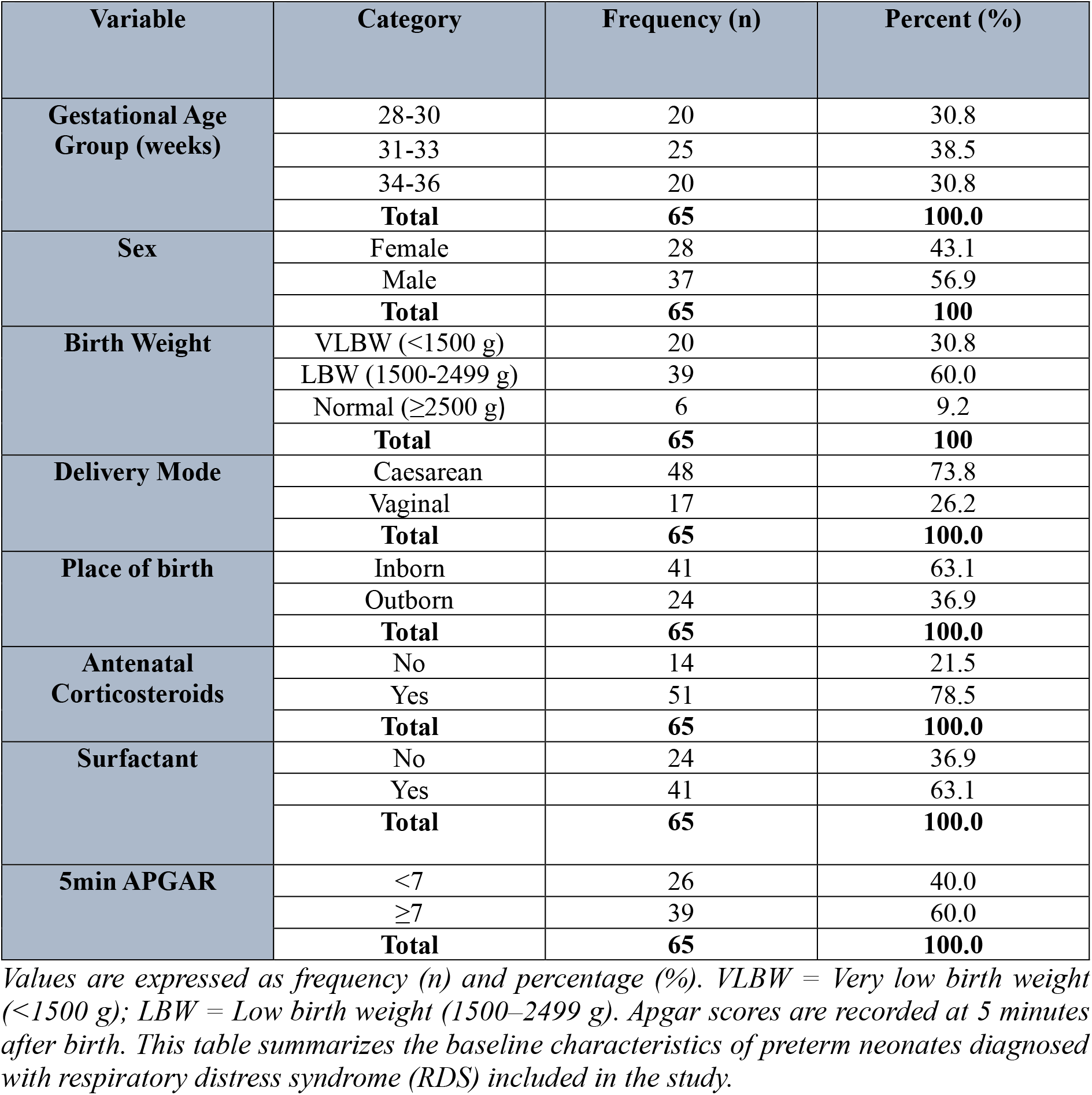
Baseline and Demographic Characteristics of Preterm Neonates with Respiratory Distress Syndrome (RDS)

Analysis of oxygen requirements and types of respiratory support across gestational age groups (combined in Table 2) revealed that younger preterm neonates required significantly higher oxygen concentrations and were more likely to need mechanical ventilation. The mean maximum FiO_2_ was 83.1 ± 18.5% in the 28–30-week group, decreasing progressively with gestational maturity (p < 0.001). This inverse relationship between gestational age and oxygen requirement is illustrated in Figure 1, whereas the distribution of respiratory support modalities (CPAP, mechanical ventilation, and nasal cannula) across gestational groups is shown separately in Figure 2. Together, these data highlight that lower gestational age was associated with higher oxygen demand and increased dependence on invasive respiratory support.

**Table 2:** Oxygen Requirement and Type of Respiratory Support Across Gestational Age Groups in Preterm Neonates with RDS.

| Gestational Age (weeks) | Frequency (n) | Maximum FiO<sub>2</sub> (%) Mean ± SD | Minimum FiO<sub>2</sub> (%) Mean ± SD | p-value (Kruskal-Wallis) | Type of Respiratory Support (n, %) | p-value (Pearson Chi-Square) |
| --- | --- | --- | --- | --- | --- | --- |
| 28-30 | 20 | 83.10 ± 18.49 | 57.05 ± 9.81 | **<0.001** | CPAP: 7 (35%), MV: 12 (60%), NC: 1 (5%) | **0.011** |
| 31-33 | 25 | 68.52 ± 20.39 | 45.24 ± 13.73 | | CPAP: 9 (36%), MV: 9 (36%), NC: 7 (28%) | |
| 34-36 | 20 | 55.60 ± 16.54 | 37.25 ± 9.84 | | CPAP: 10 (50%), MV: 2 (10%), NC: 8 (40%) | |
| **Total** | 65 | 68.17 ± 21.41 | 46.42 ± 13.79 | | CPAP: 26 (40%), MV: 23 (35.4%), NC: 16 (24.6%) | |

Values are presented as mean ± standard deviation (SD) or number (percentage).
FiO<sub>2</sub> = Fraction of inspired oxygen; CPAP = Continuous positive airway pressure; MV = Mechanical ventilation; NC = Nasal cannula.
Differences in FiO<sub>2</sub> requirements across gestational age groups were analysed using the Kruskal–Wallis test ( $p < 0.001$ ), and differences in types of respiratory support were assessed using Pearson's Chi-square test ( $p = 0.011$ ).
Lower gestational age was associated with higher oxygen requirements and greater use of mechanical ventilation, whereas older preterm neonates were more often managed with CPAP or nasal cannula.

**Figure 1.**
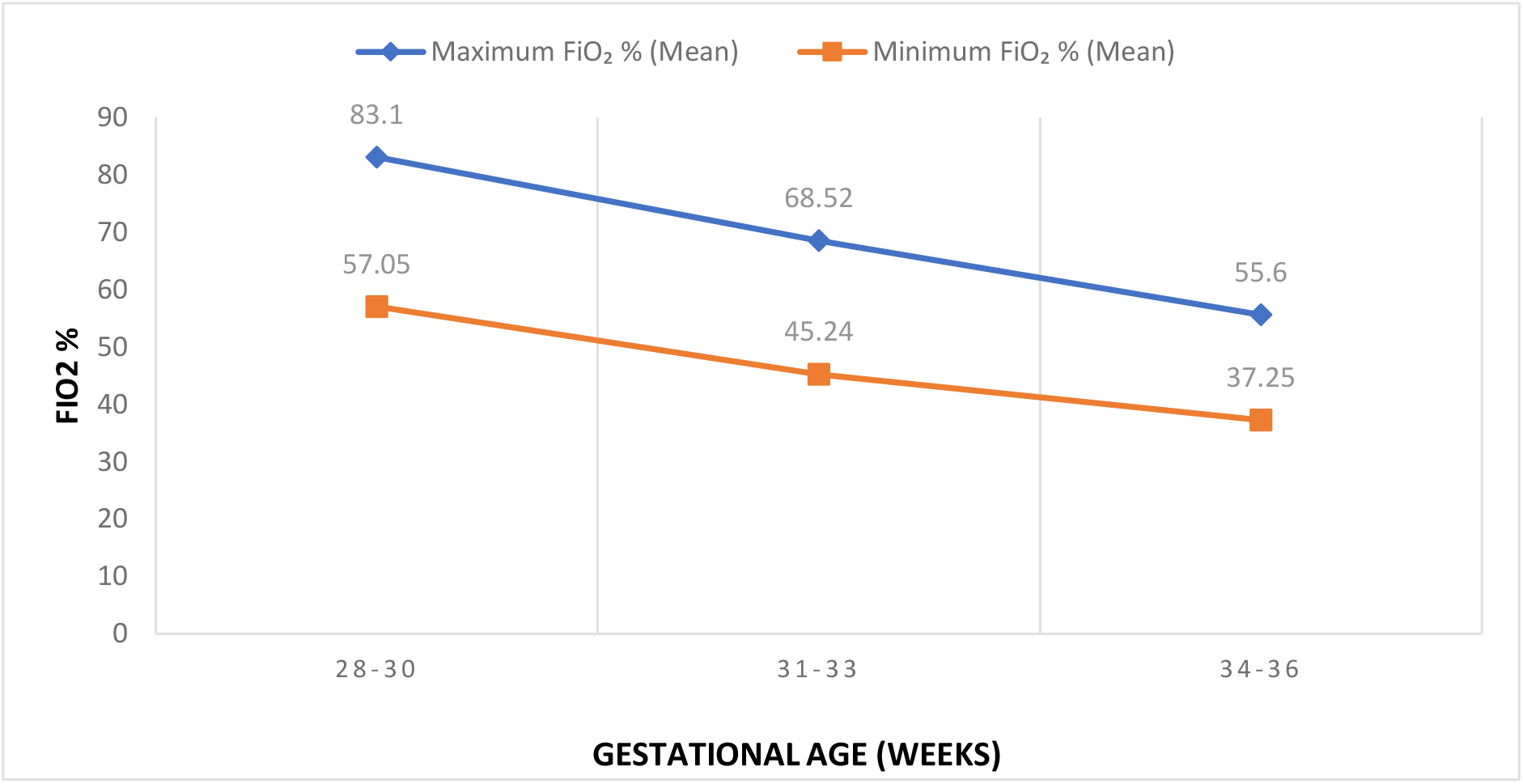
Mean FiO_2_ (%) across Gestational Age groups. The graph illustrates the mean maximum and minimum fraction of inspired oxygen (FiO_2_) across different gestational age groups (28–30, 31–33, and 34–36 weeks). Both maximum and minimum FiO_2_ levels decrease with increasing gestational age, reflecting improved respiratory maturity. The Kruskal–Wallis test was applied to compare FiO_2_ values among the groups.

**Figure 2:**
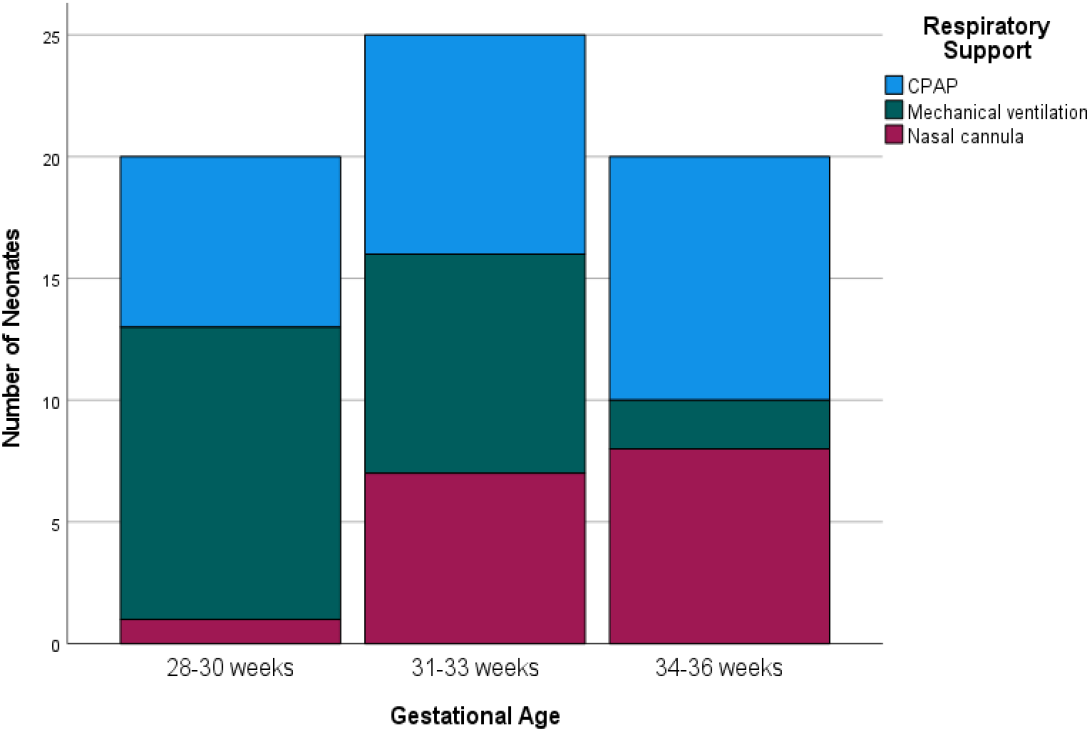
Types of Respiratory Support Across Different Gestational Ages. Distribution of respiratory support modalities among neonates by gestational age groups (28– 30 weeks, 31–33 weeks, and 34–36 weeks). Respiratory support includes continuous positive airway pressure (CPAP), mechanical ventilation, and nasal cannula. The y-axis represents the number of neonates in each category.

Baseline (pre-support) arterial blood gas (ABG) analysis demonstrated mild respiratory acidosis across all gestational groups, with mean pH values ranging between 7.22 and 7.24, as summarized in Table 3. Younger neonates had higher PaCO_2_ and lower PaO_2_ values, indicating more severe respiratory compromise. Following initiation of respiratory support, significant improvement was observed in all groups. The mean pH increased to 7.28–7.31, PaCO_2_ decreased, and both PaO_2_ and SaO_2_ improved markedly (p < 0.001 for all within-group comparisons). Despite these improvements, there were no statistically significant differences among gestational groups in the magnitude of change (ΔpH, ΔPaCO_2_, ΔPaO_2_, ΔHCO_3_^−^, or ΔSaO_2_), as shown in Table 4, suggesting similar patterns of recovery across all gestational ages.

**Table 3:** Baseline (pre-support) and stabilized (post-support) arterial blood gas (ABG) parameters by gestational age group.

| **Baseline (pre-support) ABGs by Gestational age (Group)** |  |  |  |  |  |  |
| --- | --- | --- | --- | --- | --- | --- |
| **Gestational Age (weeks)** | **Frequency (n)** | **PH pre mean ± SD** | **PaCO<sub>2</sub> pre (mmHg) mean ± SD** | **PaO<sub>2</sub> pre (mmHg) mean ± SD** | **HCO<sub>3</sub><sup>-</sup> pre (mmol/L) mean ± SD** | **SaO<sub>2</sub> pre (%) mean ± SD** |
| **28-30** | 20 | 7.22 ± 0.03 | 61.85 ± 8.31 | 37.35 ± 6.47 | 19.20 ± 2.67 | 68.72 ± 7.60 |
| **31-33** | 25 | 7.22 ± 0.05 | 55.52 ± 7.35 | 43.16 ± 7.11 | 19.08 ± 1.847 | 75.66 ± 7.98 |
| **34-36** | 20 | 7.24 ± 0.04 | 52.00 ± 6.473 | 43.50 ± 10.18 | 19.70 ± 1.89 | 75.29 ± 10.53 |
| **Stabilized (post-support) ABGs by Gestational age (Group)** |  |  |  |  |  |  |
| **Gestational Age (weeks)** | **Frequency (n)** | **PH post mean ± SD** | **PaCO<sub>2</sub> post (mmHg) mean ± SD** | **PaO<sub>2</sub> post (mmHg) mean ± SD** | **HCO<sub>3</sub> post (mmol/L) mean ± SD** | **SaO<sub>2</sub> post (%) mean ± SD** |
| **28-30** | 20 | 7.28 ± 0.03 | 53.30 ± 8.58 | 51.85 ± 6.75 | 20.35 ± 2.54 | 84.53 ± 4.06 |
| **31-33** | 25 | 7.30 ± 0.03 | 48.68 ± 6.90 | 55.96 ± 8.00 | 20.12 ± 1.79 | 87.45 ± 4.46 |
| **34-36** | 20 | 7.31 ± 0.03 | 44.95 ± 6.21 | 56.50 ± 9.25 | 21.10 ± 2.17 | 87.60 ± 4.91 |

Values are mean ± SD. PaO<sub>2</sub> = Partial pressure of oxygen; PaCO<sub>2</sub> = Partial pressure of carbon dioxide; HCO<sub>3</sub><sup>-</sup> = Bicarbonate; SaO<sub>2</sub> = Arterial oxygen saturation.

**Table 4.**
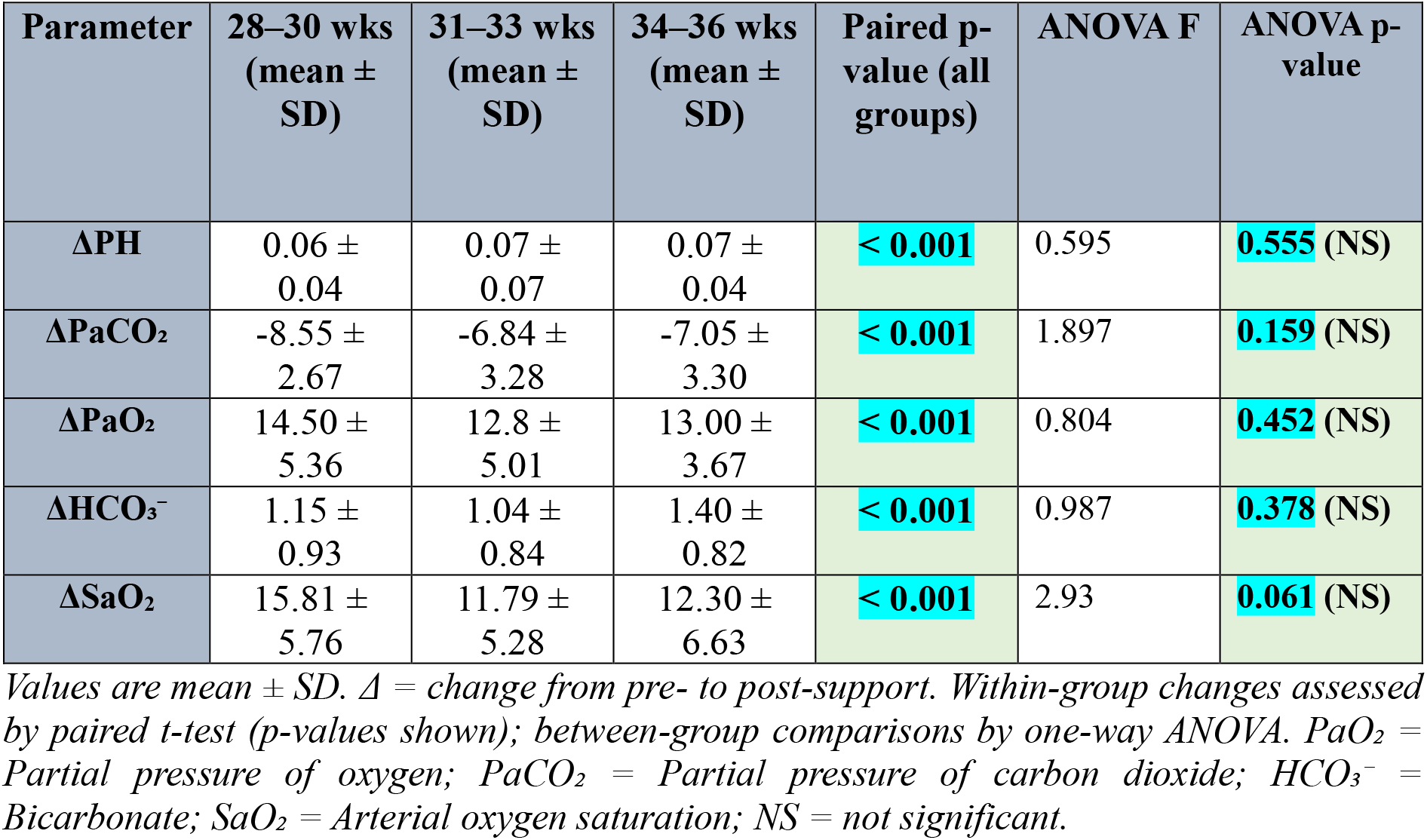
Within-group changes (Δ) in arterial blood gas (ABG) parameters and between-group comparison by gestational age.

Spearman’s correlation analysis further illustrated the relationship between gestational maturity and oxygenation parameters (Table 5). A strong negative correlation was found between gestational age and both maximum (ρ = –0.558, p < 0.001) and minimum (ρ = –0.571, p < 0.001) FiO_2_, indicating that younger preterm neonates required higher oxygen concentrations. Gestational age showed a moderate positive correlation with average pH (ρ = 0.377, p = 0.002) and weak positive correlations with PaO_2_ (ρ = 0.251, p = 0.044) and SaO_2_ (ρ = 0.274, p = 0.027). Conversely, PaCO_2_ exhibited a moderate negative correlation (ρ = –0.464, p < 0.001), while HCO_3_^−^ showed no significant association (ρ = 0.126, p = 0.316). These relationships are clearly illustrated in Supplementary Figures 1–5, which display linear trends between gestational age and individual ABG parameters.

**Table 5.** Correlation of gestational age with oxygen requirements and average arterial blood gas (ABG) parameters.

| Variable | Correlation Test | Correlation Coefficient | Sig. (2-tailed) | Interpretation |
| --- | --- | --- | --- | --- |
| Min. FiO<sub>2</sub> vs. GA | Spearman's rho | -0.571** | < 0.001 | ↑ GA → ↓ FiO<sub>2</sub><br>(Strong negative) |
| Max. FiO<sub>2</sub> vs. GA | Spearman's rho | -0.558** | < 0.001 | ↑ GA → ↓ FiO<sub>2</sub><br>(Strong negative) |
| Min. FiO<sub>2</sub> vs. Max. FiO<sub>2</sub> | Spearman's rho | +0.901** | < 0.001 | very strong positive |
| pH_avg. vs. GA | Spearman's rho | 0.377** | 0.002 | ↑ GA → ↑ pH<br>(Moderate positive) |
| PaCO<sub>2</sub> avg. vs. GA | Spearman's rho | -0.464** | < 0.001 | ↑ GA → ↓ PaCO<sub>2</sub><br>(Moderate negative) |
| PaO<sub>2</sub> avg. vs. GA | Spearman's rho | 0.251** | 0.044 | ↑ GA → ↑ PaO<sub>2</sub><br>(weak positive) |
| HCO<sub>3</sub><sup>-</sup> avg. vs. GA | Spearman's rho | 0.126** | 0.316 | No meaningful correlation |
| SaO<sub>2</sub> avg. vs. GA | Spearman's rho | 0.274** | 0.027 | ↑ GA → ↑ SaO<sub>2</sub><br>(weak positive) |

Values are Spearman's correlation coefficients ( $\rho$ ) with corresponding 2-tailed significance. "Average" values were calculated as the mean of pre- and post-support measurements. \*\* $p < 0.05$ significant; \* $p < 0.01$ highly significant. GA = Gestational Age; FiO<sub>2</sub> = Fraction of inspired oxygen; PaCO<sub>2</sub> = Partial pressure of carbon dioxide; PaO<sub>2</sub> = Partial pressure of oxygen; HCO<sub>3</sub><sup>-</sup> = Bicarbonate; SaO<sub>2</sub> = Arterial oxygen saturation

The types of respiratory support used across gestational age groups are presented in Table 2 and Figure 2. Overall, 40% of neonates were managed with continuous positive airway pressure (CPAP), 35.4% required mechanical ventilation (MV), and 24.6% were treated with nasal cannula (NC). The distribution of support types differed significantly across gestational groups (p = 0.011). The youngest neonates (28–30 weeks) were more frequently managed with mechanical ventilation, while older preterms were more often stabilized using CPAP or nasal cannula, reflecting the influence of lung maturity on respiratory support needs.

The effect of respiratory support on oxygenation status is detailed in Table 6, which shows a significant improvement in both PaO_2_ and SaO_2_ following stabilization. In the 28–30-week group, PaO_2_ increased from 37.35 ± 6.47 mmHg to 51.85 ± 6.75 mmHg, while SaO_2_ improved from 68.73 ± 7.60% to 84.53 ± 4.06%. Similar improvements were observed in the 34–36-week group, where PaO_2_ rose from 43.50 ± 10.19 mmHg to 56.50 ± 9.25 mmHg and SaO_2_ from 75.29 ± 10.53% to 87.60 ± 4.92%. Despite differences in baseline values, all neonates achieved the target saturation range of 90–94% after respiratory stabilization.

**Table 6:** Oxygen requirement and oxygenation status of preterm neonates with RDS stratified by gestational age.

| Gestational Age (weeks) | Start FiO<sub>2</sub> (%) Mean ± SD | FiO<sub>2</sub> Band (IQR range: P25–P75) | PaO<sub>2</sub> Pre (mmHg) Mean ± SD | PaO<sub>2</sub> Post (mmHg) Mean ± SD | SaO<sub>2</sub> Pre (%) Mean ± SD | SaO<sub>2</sub> Post (%) Mean ± SD | Target SaO<sub>2</sub> (%) |
| --- | --- | --- | --- | --- | --- | --- | --- |
| 28-30 | 57.05 ± 9.81 | 50-99 | 37.35 ± 6.47 | 51.85 ± 6.75 | 68.73 ± 7.60 | 84.53 ± 4.06 | 90-94 |
| 31-33 | 45.24 ± 13.73 | 32-92 | 43.16 ± 7.11 | 55.96 ± 7.99 | 75.66 ± 7.98 | 87.45 ± 4.46 | 90-94 |
| 34-36 | 37.25 ± 9.84 | 28-65 | 43.50 ± 10.19 | 55.50 ± 9.25 | 75.29 ± 10.53 | 87.60 ± 4.92 | 90-94 |

Oxygen requirement and arterial blood gas parameters in preterm neonates with respiratory distress syndrome by gestational age. Data are presented as mean ± SD unless otherwise stated; FiO<sub>2</sub> band shown as interquartile range (P25–P75). Pre = values before initiation of respiratory support; Post = values after stabilization.

Overall, the findings demonstrate that younger gestational age was consistently associated with higher oxygen requirements, more frequent need for mechanical ventilation, and greater baseline respiratory acidosis, as shown in Tables 2 and 3. However, after the initiation of appropriate respiratory support, significant improvement occurred in all groups. Correlation and graphical analyses (Table 5 and Supplementary Figures 1–5) further confirmed that advancing gestational maturity is associated with better baseline gas exchange and reduced oxygen dependency, emphasizing the physiological benefits of pulmonary development in preterm neonates with RDS.

## DISCUSSION

This study demonstrated that gestational age strongly influences respiratory management in preterm infants with respiratory distress syndrome (RDS). Neonates born at 28–30 weeks required substantially higher FiO_2_ and were more frequently managed with mechanical ventilation, whereas those at 34–36 weeks responded better to non-invasive support. These findings are consistent with earlier reports from high-resource countries, which have shown that lung immaturity and surfactant deficiency in very preterm infants increase oxygen needs and the likelihood of invasive ventilation.(3, 4, 7) However, unlike studies from high-income settings where surfactant use is routine and non-invasive support is increasingly successful, our cohort demonstrated a higher proportion of very preterm neonates requiring intubation and mechanical ventilation, reflecting differences in treatment resources and protocols in Pakistan.

Arterial blood gas parameters improved significantly after initiation of support, with increases in pH, PaO_2_, and SaO_2_ and a decline in PaCO_2_, although the magnitude of change did not differ by gestational age. This suggests that oxygen therapy is beneficial across maturity levels when titrated appropriately, in line with earlier reports.(1, 8) yet the persistence of suboptimal SaO_2_ values (<90%) across all gestational age groups contrasts with findings from centers equipped with advanced monitoring and oxygen blending devices. This highlights the practical challenges of achieving recommended saturation targets (90–94%) in resource-limited NICUs.

Our results therefore extend existing knowledge by providing the first evidence from Pakistan that gestational age not only predicts oxygen and ventilation needs but also influences the success of stabilization under conditions where surfactant, high-flow nasal cannula, and advanced monitoring are not routinely available. These insights are particularly relevant for low- and middle-income countries, where neonatal care guidelines must adapt to resource constraints.

This study has several limitations. It was conducted in a single centre with a relatively small cohort, which may limit generalizability. Only three modes of respiratory support (CPAP, mechanical ventilation, and nasal cannula) were assessed, while high-flow nasal cannula was not included. Although variables such as birth weight, antenatal steroid use, and surfactant therapy were recorded, they were not analysed in detail as potential confounders. Other important factors, such as neonatal sepsis, were also not included. In addition, long-term outcomes like bronchopulmonary dysplasia or neurodevelopment were not evaluated. As with all observational studies, residual confounding cannot be excluded. Future multicentre studies with larger samples, inclusion of additional clinical variables, and follow-up of long-term outcomes are needed to strengthen these findings.

## CONCLUSION

Gestational age is a key determinant of oxygen requirements and respiratory support in preterm neonates with RDS. Infants at earlier gestations required higher FiO_2_ and more invasive ventilation, whereas those closer to term were more often stabilized with non-invasive support. Most ABG parameters (pH, PaCO_2_, PaO_2_, SaO_2_) varied with gestational age, while HCO_3_^−^ did not, indicating that metabolic compensation was relatively uniform across groups. This study provides the first locally relevant evidence from Pakistan, demonstrating how gestational age can be used to anticipate oxygen needs and guide respiratory management in resource-limited NICUs. These findings highlight the importance of developing gestation-specific oxygen protocols to reduce the risks of both hypoxemia and hyperoxia. Future multicentre studies with larger cohorts are warranted to validate these results and inform national and regional neonatal care guidelines.

## Acknowledgment

The authors would like to thank the Neonatal Intensive Care Unit (NICU) staff of the University of Lahore Teaching Hospital, Lahore, Pakistan, for their cooperation during data collection and clinical support. The encouragement of colleagues and family members during manuscript preparation is also gratefully acknowledged.

## Author contributions

**SM:** Conceptualization, data collection, data analysis, drafting of the manuscript.

**RY:** Supervision, methodology guidance, critical revision of the manuscript.

Both authors approved the final version and are accountable for the integrity of the work.

## Data availability statement

The data that support the findings of this study are available from the corresponding author upon reasonable request. Due to patient confidentiality and institutional restrictions, the data are not publicly available.

## REFERENCES

1. Ali S, Mohammed N, Qureshi N, Gupta S. Oxygen therapy in preterm infants: recommendations for practice. Paediatrics and Child Health. 2020;31; 10.1016/j.paed.2020.10.001.

2. Bahadue FL, Soll R. Early versus delayed selective surfactant treatment for neonatal respiratory distress syndrome. Cochrane Database of Systematic Reviews. 2012(11); 10.1002/14651858.CD001456.pub2.

3. Mathias M, Chang J, Perez M, Saugstad O. Supplemental Oxygen in the Newborn: Historical Perspective and Current Trends. Antioxidants. 2021;10(12):1879; 10.3390/antiox10121879.

4. Norman M, Jonsson B, Söderling J, Björklund LJ, Håkansson S. Patterns of respiratory support by gestational age in very preterm infants. Neonatology. 2023;120(1):142–52; 10.1159/000527641.

5. Manley BJ, Arnolda GRB, Wright IMR, Owen LS, Foster JP, Huang L, et al. Nasal High-Flow Therapy for Newborn Infants in Special Care Nurseries. N Engl J Med. 2019;380(21):2031–40; 10.1056/NEJMoa1812077.

6. Abdel-Latif ME, Davis PG, Wheeler KI, De Paoli AG, Dargaville PA. Surfactant therapy via thin catheter in preterm infants with or at risk of respiratory distress syndrome. Cochrane Database Syst Rev. 2021;5(5):Cd011672; 10.1002/14651858.CD011672.pub2.

7. Abdallah Y, Mkony M, Noorani M, Moshiro R, Bakari M, Manji K. CPAP failure in the management of preterm neonates with respiratory distress syndrome where surfactant is scarce. A prospective observational study. BMC Pediatrics. 2023;23(1):211; 10.1186/s12887-023-04038-6.

8. Obst S, Herz J, Alejandre Alcazar MA, Endesfelder S, Möbius MA, Rüdiger M, et al. Perinatal Hyperoxia and Developmental Consequences on the Lung-Brain Axis. Oxidative Medicine and Cellular Longevity. 2022;2022(1):5784146; 10.1155/2022/5784146.

9. Sweet DG CV, Greisen G, et al. European Consensus Guidelines on the management of RDS – 2022 update. Neonatology. 2023;119;

10. Subramaniam P, Ho JJ, Davis PG. Prophylactic or very early initiation of continuous positive airway pressure (CPAP) for preterm infants. Cochrane Database Syst Rev. 2021;10(10):Cd001243; 10.1002/14651858.CD001243.pub4.

11. Ohuma EO, Moller A-B, Bradley E, Chakwera S, Hussain-Alkhateeb L, Lewin A, et al. National, regional, and global estimates of preterm birth in 2020, with trends from 2010: a systematic analysis. The Lancet. 2023;402(10409):1261–71; 10.1016/S0140-6736(23)00878-4.

12. Thébaud B, Goss KN, Laughon M, Whitsett JA, Abman SH, Steinhorn RH, et al. Bronchopulmonary dysplasia. Nature Reviews Disease Primers. 2019;5(1):78; 10.1038/s41572-019-0127-7.

13. McGoldrick E, Stewart F, Parker R, Dalziel SR. Antenatal corticosteroids for accelerating fetal lung maturation for women at risk of preterm birth. Cochrane Database of Systematic Reviews. 2020;2020(12); 10.1002/14651858.CD004454.pub4.

14. Hulley SB, Cummings SR, Browner WS, Grady D, Newman TB. Designing Clinical Research. 4th ed. Philadelphia: Lippincott Williams & Wilkins; 2013.

15. Kohn MA. Sample Size Calculators 2022 [Available from: https://sample-size.net.

