## Supplemental file for "Gestational Age as a Predictor of Oxygen Needs and ABG Patterns in Preterm Neonates with RDS: Evidence from a Resource-Limited NICU in Pakistan"

### Supplementary Figures

***Supp. Figure 1: Relationship Between Gestational Age and Average pH***

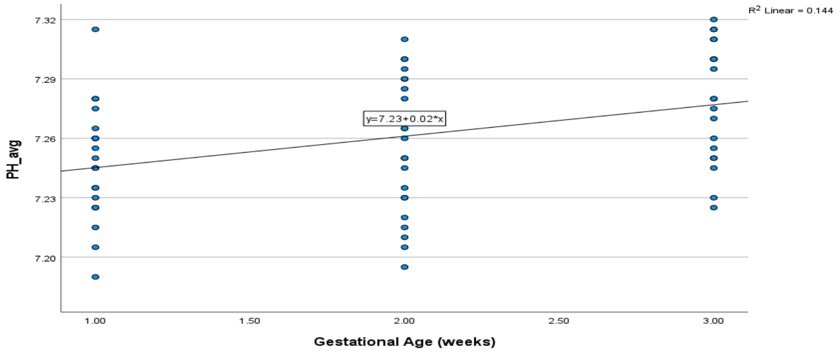


*Scatter plot depicting the association between gestational age and mean pH values, with a fitted linear regression line ($R^2$ = 0.144). Gestational age groups were coded as: 1 = 28–30 weeks, 2 = 31–33 weeks, and 3 = 34–36 weeks.*

***Supp. Figure 2: Relationship Between Gestational Age and Average $P_aCO_2$***

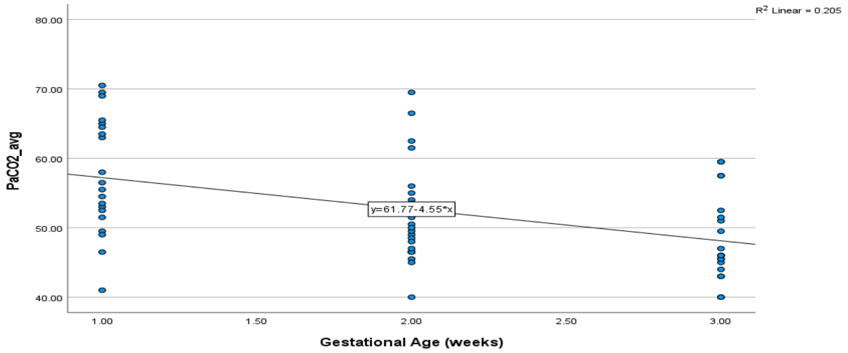


*Scatter plot depicting the association between gestational age and mean PaCO2 values, with a fitted linear regression line ($R^2$ = 0.205). Gestational age groups were coded as: 1 = 28–30 weeks, 2 = 31–33 weeks, and 3 = 34–36 weeks.*

***Supp. Figure 3: Relationship Between Gestational Age and Average $P_aO_2$***

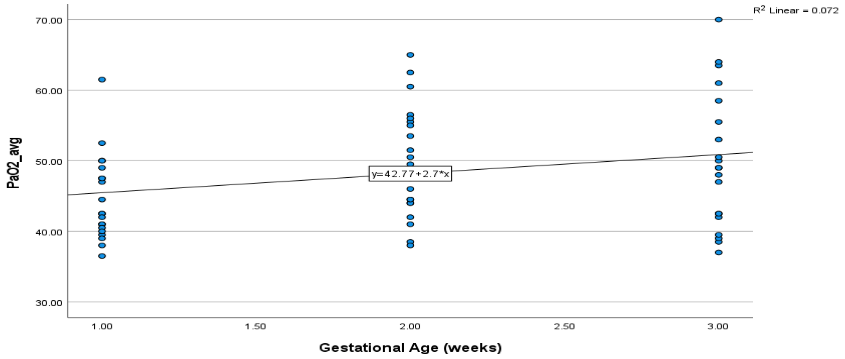


*Scatter plot depicting the association between gestational age and mean $PaO_2$ values, with a fitted linear regression line ($R^2 = 0.072$). Gestational age groups were coded as: 1 = 28–30 weeks, 2 = 31–33 weeks, and 3 = 34–36 weeks.*

***Supp. Figure 4: Relationship Between Gestational Age and Average $HCO_3^-$***

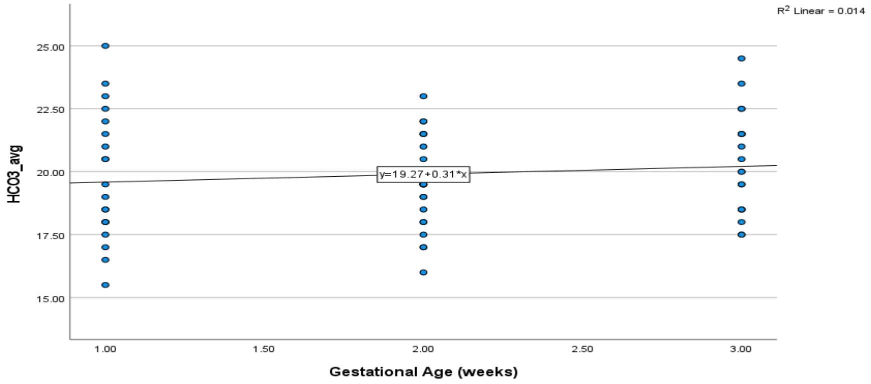


*Scatter plot depicting the association between gestational age and mean $HCO_3^-$ values, with a fitted linear regression line ($R^2 = 0.014$). Gestational age groups were coded as: 1 = 28–30 weeks, 2 = 31–33 weeks, and 3 = 34–36 weeks.*

***Supp. Figure 5: Relationship Between Gestational Age and Average $S_aO_2$***

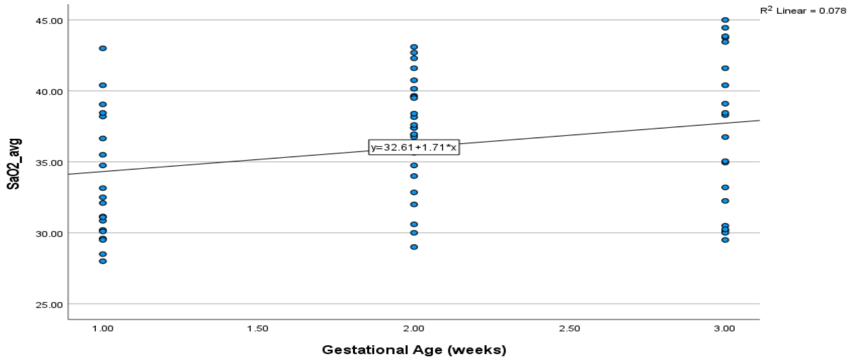


*Scatter plot depicting the association between gestational age and mean $SaO_2$ values, with a fitted linear regression line ($R^2 = 0.078$). Gestational age groups were coded as: 1 = 28–30 weeks, 2 = 31–33 weeks, and 3 = 34–36 weeks.*

**QUESTIONNAIRE**

#### Section A: Neonatal Demographics

1. Study ID: ____________

2. Gestational age (in weeks): ______ weeks

3. Birth weight (in grams): ______ grams

4. Gender: ☐ Male ☐ Female

5. Mode of delivery: ☐ Vaginal ☐ Cesarean ☐ Not captured

6. Place of Birth: ☐ Inborn ☐ Out born

7. APGAR score at 5 minutes: ______ /10

#### Section B: Maternal and Perinatal History

8. Antenatal steroid given? ☐ Yes ☐ No

9. Number of steroid doses received:

   ☐ 1 dose ☐ 2 doses ☐ Unknown

10. Any maternal infection (e.g., chorioamnionitis)? ☐ Yes ☐ No

11. Hypertension or pre-eclampsia? ☐ Yes ☐ No

#### Section C: RDS and Respiratory Support

12. Diagnosed with RDS? ☐ Yes ☐ No

13. Chest X-ray consistent with RDS? ☐ Yes ☐ No

14. Silverman Anderson score: ______ /10

15. RDS severity: ☐ Mild ☐ Moderate ☐ Severe

16. Received surfactant? ☐ Yes ☐ No

17. Number of surfactant doses: ☐ 1 ☐ 2 ☐ >2

18. Type of oxygen support:

☐ Nasal cannula

☐ CPAP

☐ HFNC

☐ Mechanical ventilation

19. Minimum $FiO_2$ (%): _____%

20. Maximum $FiO_2$ (%): _____%

#### Section D: ABG Parameters

| **Variable** | **Pre-Respiratory Support ABG** | **Post-Respiratory Support ABG** |
|---|---|---|
| pH | | |
| $PaO_2$ (mmHg) | | |
| $PaCO_2$ (mmHg) | | |
| $HCO_3^-$ (mmol/L) | | |
| SaO2 | | |

#### Section E: Outcome

26. Length of NICU stay (days): ______ days

27. Discharged alive? ☐ Yes ☐ No

28. Complications?

☐ PDA ☐ IVH ☐ BPD ☐ Sepsis ☐ NEC ☐ None

### Permission letter

#### Department of health Professional Technologies

FACULTY OF ALLIED HEALTH SCIENCES, UNIVERSITY OF LAHORE.

##### Title Approval Form

| | |
|---|---|
| **Student's name** | Shoaib Mehmood |
| **Supervisor's name** | Ms. Ruhamah Yousaf |
| **Program** | Bachelor of Science in Respiratory Therapy |
| **Session** | 2022-2026 |

| | |
|---|---|
| **Title of study** | Gestational Age as a Predictor of Oxygen Needs and ABG Patterns in Preterm Neonates with RDS: Evidence from a Resource-Limited NICU in Pakistan |
| **Objective (s) of the study** | 1. To identify and compare the oxygen needs of pre-mature neonates who are diagnosed with respiratory distress syndrome (RDS) at various gestational ages.<br>2. To assess and compare arterial blood gas (ABG) parameters (PH, PaO 2, PaCO 2, HCO3) in preterm neonates with RDS of different gestation ages.<br>3. To evaluate the relationship between gestational age, oxygen requirement and ABG parameters among preterm neonates who have RDS.<br>4. To determine typical oxygen delivery methods in gestations of varying age groups. |
| **Methodology**<br><br>**(Design, sample size, sampling technique, Study setting, inclusion and exclusion criteria and Tool)** | **Study Design:** Analytical cross-sectional study<br>**Sample Size:** 62 neonates<br>**Sampling Technique:** Non-probability sampling technique<br>**Study Setting:** NICU setting (Children hospital Lahore) |

| Inclusion Criteria | Exclusion Criteria |
|---|---|
| Preterm neonates (<37 gestational weeks) | Neonates that have significant congenital abnormality |
| Diagnosed with RDS according to the clinical and radiographic evidence | Neonates whose major diagnosis is given as birth asphyxia or sepsis |
| Admitted in the NICU within the first 24 hours after birth | Incomplete or missing ABGs data |
| Requiring oxygen therapy | |
| Available ABG analysis within 6 hours of admission | |

| | |
|---|---|
| **Data collection procedure** | Procedure will include obtaining following NICU parameters of preterm neonates:<br>• Demographic information (gestation age, weight at birth, sex)<br>• RDS clinical diagnosis<br>• Type and duration of oxygen therapy<br>• Oxygenation method<br>• Parameters such as (PH, PaO2, PaCO2 and HCO3-) in arterial blood gas (ABGs) |
| **Ethical considerations** | • Approval will be taken from hospital after taking request form from the university department.<br>• A written informed consent by the parents /guardians will be taken prior to the collection of data.<br>• Information security will be with strict sense of confidentiality and privacy.<br>• There will be no intervention, but only observational data will be measured. |
| **Any specific point** | Aims is to look O2 trends gestational wise will be measured and ABGs profile in improving individualized Respiratory care of preterm neonates with RDS |
| **Impact of this research on society** | The study will improve neonatal care by guiding O2 therapy and ABGs monitoring based on gestational age, reducing complications and Optimizing outcomes in preterm neonates with RDS |
| **Remarks of committee** | |
| **Signature of the committee:** | |

Dr. Sumbal Shahbaz
Assistant professor / Head of Department
[illegible] (Hons) (Anaesthesia), M.Phil (P.H), PHd (P.H)
Department of Health Professional Technologies (DHPT)
Faculty of Allied Health Sciences
The University of Lahore, Pakistan

THE UNIVERSITY OF LAHORE
A Project of Ibadat Educational Trust

**Date: 02/06/2025** **Centre/Admin FAHS/195/25**

**To,**

**Medical Superintendent,**

**The University of Lahore Teaching Hospital (UOLTH), Lahore.**

Subject: **Request for Permission to Collect Research Data**

Respected Sir/Mam,

It is stated that **Mr. Shoaib Mehmood** with **Reg. No: 70133025** is a bonafide student of Bachelor of Science in Respiratory Therapy in Department of Health Professional Technologies, Faculty of Allied Health Sciences, The University of Lahore.

His research is going to be start on the topic **"Gestational Age as a Predictor of Oxygen Needs and ABG Patterns in Preterm Neonates with RDS: Evidence from a Resource-Limited NICU in Pakistan."**

You are requested to accommodate him in the respective Department/ward of your esteemed hospital to collect research data.

**Dr. Sumbal Shahbaz**
Head of Department
Health Professional Technology
The University of Lahore.


NS
19/7 Permitted



**STROBE Checklist for Cross-Sectional Study**

| Section / Topic | Item No. | Recommendation | Reported on Page | Notes / Correction |
|---|---|---|---|---|
| Title & Abstract | 1a | Indicate the study design in the title or abstract | 1–2 | ✅ Title and abstract clearly identify the study as a cross-sectional study conducted in a NICU. |
| | 1b | Provide an informative and balanced abstract summarizing objectives, methods, results, and conclusions | 2 | ✅ Structured abstract with background, objective, methods, results, and conclusion. |
| Introduction | 2 | Explain the scientific background and rationale for the study | 3–5 | ✅ Clearly explains RDS pathophysiology, oxygen therapy, and research gap in Pakistan. |
| | 3 | State specific objectives, including any prespecified hypotheses | 5 | ✅ Objective explicitly stated at the end of the introduction. |
| Methods | 4 | Present key elements of study design early in the paper | 6 | ✅ Cross-sectional analytical study described under "Study Design and Setting." |
| | 5 | Describe the setting, locations, | 6 | ✅ Conducted in NICU, University of Lahore Teaching |

| | | | | |
|---|---|---|---|---|
| | | and relevant dates | | Hospital, Pakistan, over four months. |
| | 6 | Give eligibility criteria and sources/methods of participant selection | 6 | ✅ Inclusion and exclusion criteria (preterm neonates with RDS, <37 weeks, no congenital anomalies). |
| | 7 | Clearly define all variables, including outcomes, exposures, confounders | 7 | ✅ Independent: Gestational age. Dependent: $FiO_2$, ABG parameters, and type of respiratory support. |
| | 8 | For each variable, provide data sources and measurement methods | 7 | ✅ Data extracted from NICU records; pre- and post-support ABGs measured by standard equipment. |
| | 9 | Describe efforts to address potential sources of bias | 7 | ✅ Standardized data collection forms used to minimize measurement bias. |
| | 10 | Explain how the study size was arrived at | 6 | ✅ Sample size calculated using correlation-based formula; total 65 |

| | | | | |
|---|---|---|---|---|
| | | | | neonates enrolled. |
| | 11 | Explain how quantitative variables were handled in the analyses | 7 | ✅ Continuous variables as mean ± SD; categorical as n (%); appropriate statistical tests used. |
| | 12a | Describe all statistical methods, including those used to control for confounding | 8 | ✅ ANOVA, Kruskal–Wallis, Chi-square, paired t-test, Spearman's correlation applied. No confounder adjustment required. |
| | 12b | Describe any methods used to examine subgroups and interactions | 8 | ✅ Gestational age subgroups analyzed (28–30, 31–33, 34–36 weeks). |
| | 12c | Explain how missing data were addressed | – | ✅ No missing data encountered for primary variables. |
| | 12d | If applicable, explain how loss to follow-up was addressed | – | N/A – Cross-sectional study (no follow-up). |
| | 12e | Describe any sensitivity analyses | – | N/A – Sensitivity analysis not applicable. |

| | | | | |
|---|---|---|---|---|
| Results | 13a | Report numbers of individuals at each stage of the study | 9, Table 1 | ✅ Participant distribution by gestational age, sex, and weight reported. |
| | 13b | Give reasons for non-participation at each stage | – | ✅ All eligible neonates were included; no post-enrolment exclusions. |
| | 13c | Consider use of a flow diagram | – | Optional – not required for single-centre dataset. |
| | 14a | Give characteristics of study participants | 10–11, Table 1 | ✅ Baseline and demographic characteristics detailed. |
| | 14b | Indicate number of participants with missing data for each variable | – | ✅ No missing data for key outcomes. |
| | 14c | Summarize follow-up time (if applicable) | – | N/A – Single-timepoint assessment. |
| | 15 | Report numbers of outcome events or summary measures | 12–17, Tables 2–5 | ✅ $FiO_2$, ABG, and support type outcomes reported. |
| | 16a | Give unadjusted estimates and precision | 12–17 | ✅ Unadjusted comparisons across GA groups with p-values. |

| | | | | |
|---|---|---|---|---|
| | 16b | Report category boundaries when continuous variables categorized | 9 | ✅ Gestational age groups (28–30, 31–33, 34–36 weeks) defined. |
| | 17 | Report other analyses done (e.g., subgroup or sensitivity analyses) | 15–16 | ✅ Correlation and subgroup analyses performed; no sensitivity analysis applicable. |
| Discussion | 18 | Summarize key results with reference to study objectives | 18 | ✅ Results summarized at start of discussion. |
| | 19 | Discuss limitations of the study | 20 | ✅ Limitations discussed (sample size, single-centre, lack of long-term outcomes). |
| | 20 | Give cautious interpretation of results, considering objectives and other studies | 18–21 | ✅ Compared findings with previous literature; contextualized to resource-limited NICUs. |
| | 21 | Discuss generalisability (external validity) of the study results | 21 | ✅ Findings applicable to low-resource and LMIC NICU settings. |

| | | | | |
|---|---|---|---|---|
| Other Information | 22 | Give source of funding and role of funders | 22 | ✅ No funding received. |
| | 23 | Provide complete reference list | 23–24 | ✅ References (1–15) provided; |
